# Current Status and Emerging Trends of COVID-19-related Studies in Seven ‘Tropical Medicine’-entitled Journals

**DOI:** 10.1101/2023.01.30.23285173

**Authors:** Xuejuan Zhang, Ziqiao Zhong, Peili Luo, Chune Zhu, Ying Huang, Chuanbin Wu, Xin Pan, Zhengwei Huang

## Abstract

**Background:** Coronavirus disease 2019 (COVID-19), caused by severe acute respiratory syndrome coronavirus 2 (SARS-CoV-2), has created enormous medical and economic burdens on human society. However, the co-existence of COVID-19 and diseases in tropical regions is not taken seriously. To improve the understanding of the current status and trends on crosstalk of COVID-19 and tropical diseases, this paper provided an analysis, from a bibliometric perspective, of the COVID-19-related publications in ‘Tropical Medicine’-entitled journals.

**Methods:** We used Clarivate Analytics and VOSviewer to analyze 783 publications in seven ‘Tropical Medicine’-entitled journals. Document overview, basic bibliometric characteristics, citation performance, co-authorship, co-citation, bibliographic coupling, and co-occurrence of keywords and terms were summarized in this article.

**Results:** Document overview revealed that 76.12% of the related publications were published in open access mode, and basic bibliometric characteristics indicated that the year 2021 was the peak of the number of publications, the documents in the seven journals were unevenly distributed, and ‘article’ was the main publication type. The citation performance analysis elucidated that the documents of interest were frequently cited. The co-authorship analysis showed cooperation networks on the level of region, organization and author. General knowledge of COVID-19 was the overlap of co-citation and bibliographic coupling behavior. Finally, the co-occurrence of keywords and terms revealed the current and emerging hotspots.

**Conclusions:** The main current research focuses in ‘Tropical Medicine’-entitled journals are the clinical features of COVID-19 patients, and the emerging trends are the hesitancy in making vaccines against SARS-CoV-2 and the circumstance where COVID-19 coexisted with tropical diseases. In summary, this bibliometric analysis of COVID-19-related studies in seven ‘Tropical Medicine’-entitled journals highlights the current research focuses of this field to inspire future studies.

## 1 Introduction

Coronavirus Disease 2019 (COVID-19), induced by Severe Acute Respiratory Syndrome Coronavirus 2 (SARS-CoV-2)^[1]^, has become a severe worldwide epidemic. As an indispensable part of the world, the tropical region (defined by the region between Tropic of Capricorn and Tropic of Cancer) is inevitably impacted by the COVID-19 epidemic^[2]^. According to the data of World Health Organization COVID-19 dashboard (https://covid19.who.int/), there are 9138808 cumulative confirmed cases and 173674 cumulative death cases by 6:19 pm (CEST), July 6^th^, 2022. Besides, the co-existence of COVID-19 and diseases in tropical regions, especially neglected tropical diseases, makes the pandemic even more threatening and complicated^[3]^.

In this scenario, scientific journals within the scope of tropical medicine have paid great attention to the evolving state of the COVID-19 pandemic. Seven ‘Tropical Medicine’-entitled journals indexed by Science Citation Index Expanded in Web of Science Core Collection, i.e., *Journal of Tropical Medicine, Southeast Asian Journal of Tropical Medicine and Public Health, Tropical Medicine & International Health, Tropical Medicine and Infectious Disease, Transactions of the Royal Society of Tropical Medicine and Hygiene, Asian Pacific Journal of Tropical Medicine* and *American Journal of Tropical Medicine and Hygiene*, must be mentioned. A considerable number of valuable contributions regarding COVID-19 have been published by the seven journals. The detailed information about these journals can be found in **Table 1**.

**Table 1.**
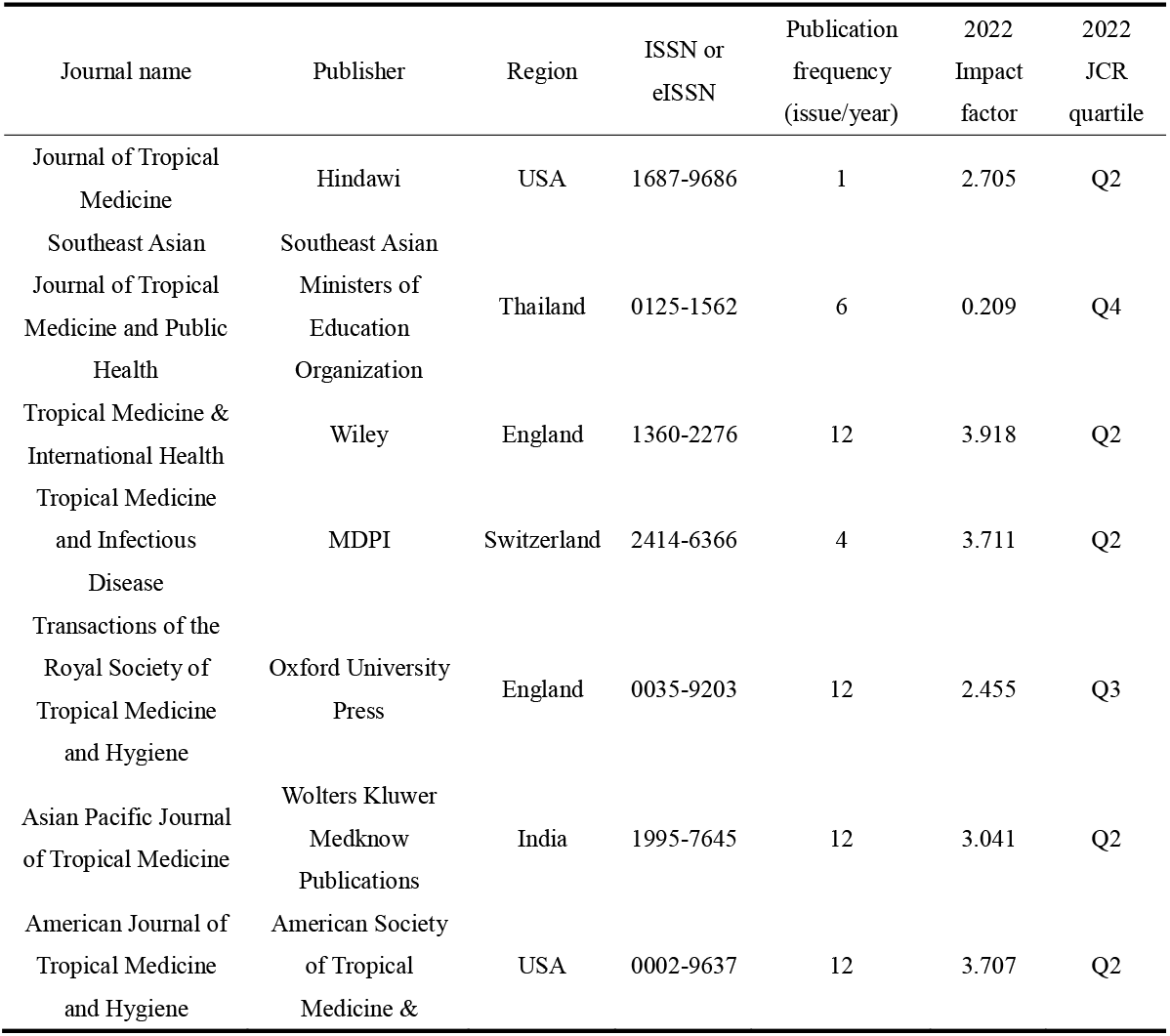
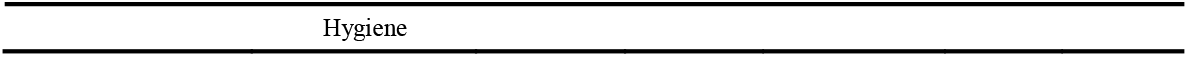
Information about the seven ‘Tropical Medicine’-entitled journals. Collected from journal official websites and 2022 Clarivate Journal Citation Reports (JCR).

A question raises ‘what are the current status and emerging trends about the relevant studies?’ To address this issue, bibliometric analysis should be performed^[4]^. Although some general bibliometric research upon COVID-19 has been accomplished^[5-9]^, by far, no specific bibliometric study on the COVID-19-related publications in ‘Tropical Medicine’-entitled journals, which impedes the understanding of the current status and emerging trends.

In this work, a comprehensive bibliometric study was carried out. Using Clarivate Analytics and VOSviewer as tools, document overview, basic bibliometric characteristics, citation performance, co-authorship, co-citation, bibliographic coupling, and co-occurrence of keywords and terms were analyzed thoroughly. The current status and emerging trends were further summarized based on the results. We hoped this piece of work could help to master the *status quo* of COVID-19-related studies in important ‘Tropical Medicine’-entitled journals and trigger the upcoming contributions.

## 2 Methods

The bibliometric methodology of this work is summarized in **Scheme 1**.

**Scheme 1.**
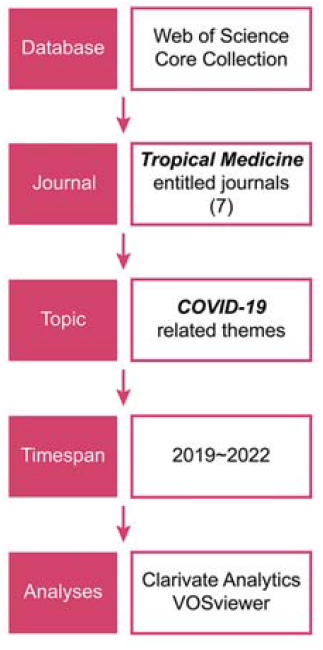
Framework of bibliometric methodology.

### 2.1 Document survey

The COVID-19-related publications in seven ‘Tropical Medicine’-entitled journals, viz. *Journal of Tropical Medicine, Southeast Asian Journal of Tropical Medicine and Public Health, Tropical Medicine & International Health, Tropical Medicine and Infectious Disease, Transactions of the Royal Society of Tropical Medicine and Hygiene, Asian Pacific Journal of Tropical Medicine*, and *American Journal of Tropical Medicine and Hygiene* were retrieved from Web of Science Core Collection database, accessed through Jinan University.

Document survey was performed at 2 pm, on June 30^th^, 2022 (time zone: UST + 8). The full query set was as follows. Definitions of codes: SO-journal title, TS-topic. ‘OR’ and ‘AND’ were Boolean operators. #1: (SO = Journal of Tropical Medicine) OR (SO = Southeast Asian Journal of Tropical Medicine and Public Health) OR (SO = Tropical Medicine & International Health) OR (SO = Tropical Medicine and Infectious Disease) OR (SO = Transactions of the Royal Society of Tropical Medicine and Hygiene) OR (SO = Asian Pacific Journal of Tropical Medicine) OR (SO = American Journal of Tropical Medicine and Hygiene).

#2: (TS = COVID) OR (TS = COVID 19) OR (TS = COVID-19) OR (TS = COVID 2019) OR (TS = Coronavirus Disease 2019) OR (TS = COVID-2019) OR (TS = SARS CoV 2) OR (TS = SARS-CoV-2) OR (TS = Severe Acute Respiratory Syndrome Coronavirus 2) OR (TS = SARS Coronavirus 2) OR (TS = 2019-nCoV) OR (TS = 2019 nCoV).

#3: #1 AND #2.

Further, considering the outbreak time of COVID-19, the publication time was refrained to 2019∼2022, by 2 pm, June 30^th^, 2022 (time zone: UST + 8). The retrieved documents were exported as plain text and tab-delimitated files for subsequent analyses.

### 2.2 Bibliometric analysis

The documents were analyzed by the analytical module of Clarivate Analytics (associated with Web of Science) ^[10]^, and VOSviewer (version 1.6.17, Leiden University, Leiden, Netherlands)^[11]^, to showcase the knowledge domain and emerging trends.

Clarivate Analytics: Information about language, open access modes, publication years, journal titles, research areas, publication types, geographic distribution, organizations, authors, and funding agencies was analyzed. Moreover, citation performance was demonstrated by total citing articles, citing articles without self-citations, average citing articles per item, total times cited, times cited without self-citations, average times cited per item, and Hirsch index (*H*-index). The top-20 cited publications were extracted.

VOSviewer: Networks about co-authorship (regions, organizations and authors), co-citation, bibliographic coupling and co-occurrence (author keywords + keywords plus and title/abstract terms) were created. The full counting method was utilized to fabricate the clustered or time-overlay networks. Threshold values for network preparation: co-authorship, regions-15; co-authorship, organizations-10; co-authorship, authors-5; co-citation-20; bibliographic coupling-30; co-occurrence, author keywords + keywords plus-5; co-occurrence, title/abstract terms-50.

## 3 Results

### 3.1 Document overview

A literature survey returned 783 documents in total. All (783, 100%) of these documents were written in English, which favored global communications^[12]^. A large proportion (596, 76.12%) of them were published in open access (OA) mode, which demonstrated a relatively high level in a research field compared to previous studies^[13-15]^. The OA mode made it free to download and read the published COVID-19-related works, increasing the accessibility and visibility^[16]^, which might be especially beneficial to developing regions.

### 3.2 Basic bibliometric characteristics

The distribution of publication years of the 783 documents is depicted in **Figure 1(a)**. There were 223 (28.48%), 463 (59.13%) and 97 (12.39%) documents published in 2020, 2021 and 2022, respectively. ‘Tropical Medicine’-entitled journals showed their concerns about COVID-19 in an early stage of the epidemic (the year 2020), and the first five papers were published in March 2020^[17-21]^. The year 2021 witnessed a 107.62% increase in publication numbers to 2020, suggesting the raising concerns.

**Figure 1.**
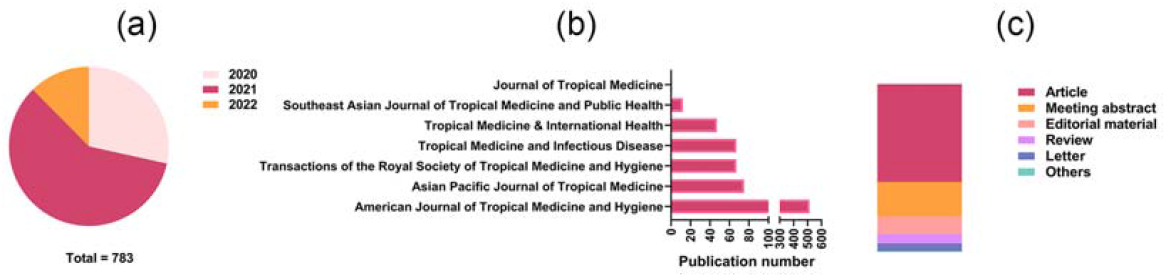
Distribution of publication years (a), journals (b) and publication types (c) of the retrieved documents.

**Figure 1(b)** shows the distribution of journals. All seven journals of interest had published relevant works, but the publication number varied. *American Journal of Tropical Medicine and Hygiene* published most papers (514, 65.65%), followed by *Asian Pacific Journal of Tropical Medicine* (75, 9.58%), *Transactions of the Royal Society of Tropical Medicine and Hygiene* (67, 8.56%) and *Tropical Medicine and Infectious Disease* (67, 8.56%). The other journals published less than 50 related works: *Tropical Medicine & International Health* (47, 6.00%), *Southeast Asian Journal of Tropical Medicine and Public Health* (12, 1.53%) and *Journal of Tropical Medicine* (1, 0.13%). It should be noted that the varied numbers only partly reflexed the emphases laid on COVID-19 research, but were not indicators of the journal’s scientific standard.

The distribution of publication types is summarized in **Figure 1(c)**. Articles (456, 58.24%) were the major publication type. Interestingly, meeting abstracts accounted for a considerable proportion of 20.69%. It was inferred that some important conferences were held to review the *status quo* of the epidemic and the state-of-the-art of therapeutic strategies, and the accepted abstracts of these meetings might provide meaningful cutting-edge information. The percentage of reviews was relatively low (5.62%), which was lower than editorial materials (10.35%) and close to letters (4.73%). It was anticipated that more reviews could be written to summarize the current status of this field, which would help to figure out the future directions^[22]^.

**Figure 2(a)** depicts the top geographical distribution of the retrieved documents. Twelve regions contributed at least 30 publications. The USA was the main contributor, with 281 publications, followed by England with 117 publications. The other regions (India, Nigeria, Brazil, Thailand, Netherlands, China, Pakistan, Bangladesh, Ethiopia and Switzerland) contributed less than 100 publications. It was shown that both topical and non-tropical, developed, and developing regions participated in the contribution. Hence, it was a global trend to publish COVID-19-related works in ‘Tropical Medicine’-entitled journals.

**Figure 2.**
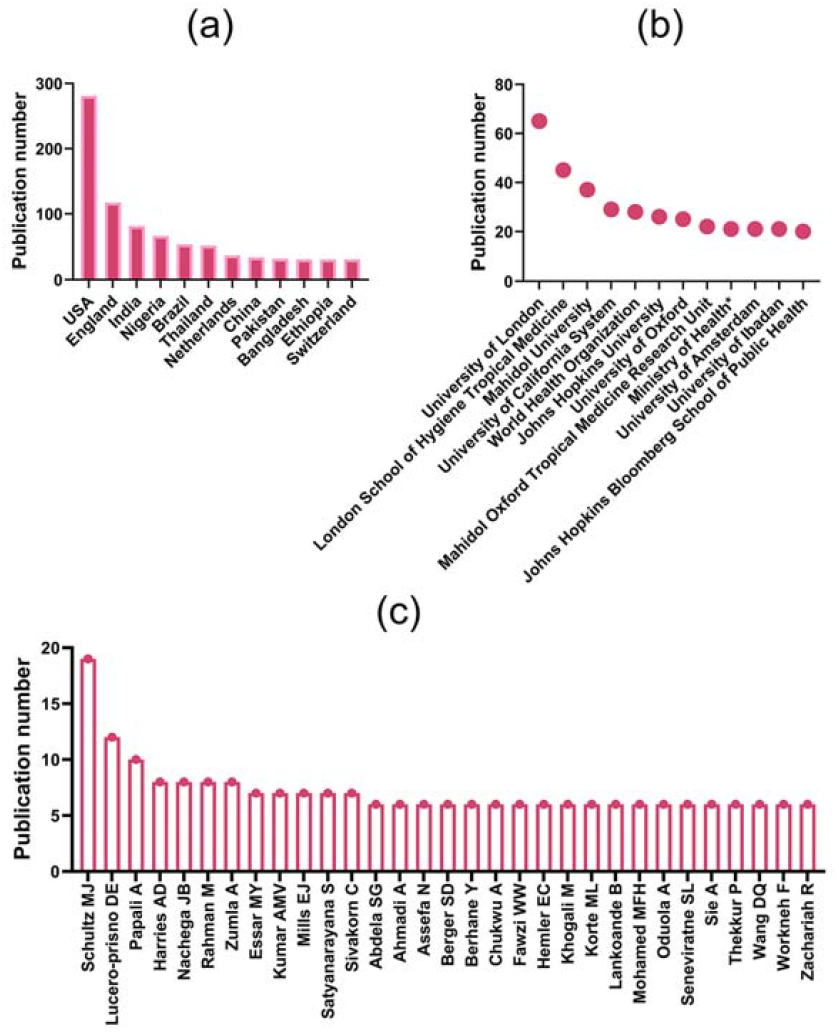
Top geographical distribution (a), distribution of top organizations (b) and top authors (c) of the retrieved documents. Of note, ‘Ministry of Health’ in (b) included Ministry of Health of Guinea (1), Bhutan (1), Vietnam (1), Brazil (2), Kenya (1), Brunei (1), Saudi Arabia (2), Uganda (4), Italy (1), Sri Lanka (1), Dem Rep Congo (2), Malawi (1), Nicaragua (1), Somalia (1), Rwanda (1) and Israel (1).

According to **Figure 2(b)**, 11 organizations contributed ≥ 20 publications; ‘Ministry of Health’ ranking the 9^th^ was not an individual organization, but a combination of the ministries in different regions (Guinea, Bhutan, Vietnam, Brazil, Kenya, Brunei, Saudi Arabia, Uganda, Italy, Sri Lanka, Dem Rep Congo, Malawi, Nicaragua, Somalia, Rwanda and Israel, most of them were tropical regions). As institutes in tropical regions, Mahidol University (Thailand), Mahidol-Oxford Tropical Medicine Research Unit (Thailand), and University of Ibadan (Nigeria) appeared on the list of top contributors. University of London (England) and London School of Hygiene & Tropical Medicine (England) acted as the top organizations, and the University of Oxford (England), University of Amsterdam (Netherlands), *etc*. also contributed a large number of publications, revealing that institutions in non-tropical regions were concerned about the crosstalk of COVID-19 and tropical diseases as well. World Health Organization to improve the health level of all humankind ranked as top 5^[23]^, which elaborated its concerns about the impact of COVID-19 in tropical regions. Additionally, the geographical distribution of the top organizations was basically in parallel with **Figure 2(a)**.

The most fruitful authors (31 with over 5 publications) in this field are listed in **Figure 2(c)**. Schultz MJ (primary affiliation: Mahidol University, Thailand), Lucero-prisno DE (primary affiliation: London School of Hygiene & Tropical Medicine, England), and Papali A (Atrium Health, USA) with 19, 12 and 10 papers were the top-3 authors, who were all from the top contributing regions in **Figure 2(a)**. The other authors published 6∼8 papers. For newly emerging investigators, it would be advisable to seek collaboration with these fruitful authors.

The basic bibliometric characteristics of 783 documents were described above. A peak number of publications occurred in the year 2021, the documents were distributed unevenly in the seven journals and ‘article’ was the major publication type. The top contributing regions, organizations and authors were summarized, and the results showed that global discussion was included in the seven journals. Some peripheral data can be found in **Supplementary Material Tab. S1** and **S2**.

### 3.3 Citation performance

The citation performance of the 783 documents was scrutinized, and the results are displayed in **Figure 3**. The numbers of total citing articles, citing articles without self-citations, total times cited and times cited without self-citations were 5372, 5195, 6015 and 5747, respectively. It was then calculated that the self-citation only occupied a proportion of approximately 4%, which was a quite low rate compared to published literature ^[24-26]^. It was considered that with a lower self-citation rate, the citation performance would be more significant and convincing^[27]^. The average number of citing articles and times cited per item were 6.86 and 7.68, respectively. Furthermore, the *H*-index was 34, meaning that 34 publications were cited at least 34 times. Compared with some bibliometric studies of COVID-19^[28-30]^, this *H*-index was moderate. Therefore, the citation performance could be further improved, and more attention from the science community was needed. It was anticipated that the COVID-19-related research published in ‘Tropical Medicine’-entitled journals could provide more insights into the COVID-19 management during the post-COVID-19 era, by serving as valuable references.

**Figure 3.**
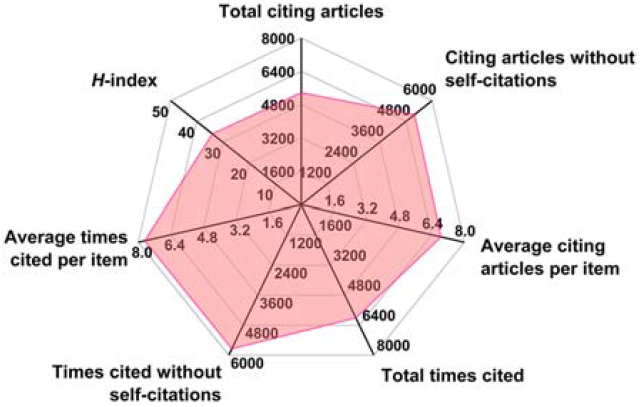
Radar map of citation performance of the retrieved documents.

The top-20 most cited papers were extracted, listed in **Table 2**. Documents published in *Tropical Medicine & International Health, American Journal of Tropical Medicine and Hygiene, Asian Pacific Journal of Tropical Medicine, Transactions of the Royal Society of Tropical Medicine and Hygiene*, and *Tropical Medicine and Infectious Disease* (5 out of 7 journals) were involved in the list. Correspondingly, 1, 14, 2, 1, 1 and 1 papers were published in the above journals. The journal *American Journal of Tropical Medicine and Hygiene* with most entrances was also the top 1 publishing journal in **Figure 1(b)**. It was perceived that this journal might be one of the flagship journals in its area. No entrance in **Table 2** was published in *Southeast Asian Journal of Tropical Medicine and Public Health* and *Journal of Tropical Medicine*, probably owing to the small number of relevant publications in these journals. All entrances were published in 2020 and could be regarded as pioneer research. The distribution types were to some degree interesting: 3 were editorial materials, 3 were reviews and 14 were articles. The ratio was similar to that of **Figure 1(c)**. Editorial materials accounted for a certain proportion, and even the top 1 paper was an editorial material^[18]^. The appearance of editorial materials in the most cited papers was a rare case in bibliometric analyses^[31-33]^. It should be explained by that the editorial members of the related journals were highly concerned about the COVID-19 epidemic, and they offered useful insights that deserved widespread attention (especially regarding the evolving status of the epidemic^[18, 34]^, and the administration of potential SARS-CoV-2 inhibitors)^[35]^. In addition, the total citations per paper varied between 52 and 930, 7∼132-fold higher than the average times cited obtained in **Figure 3** (∼7).

**Table 2.**
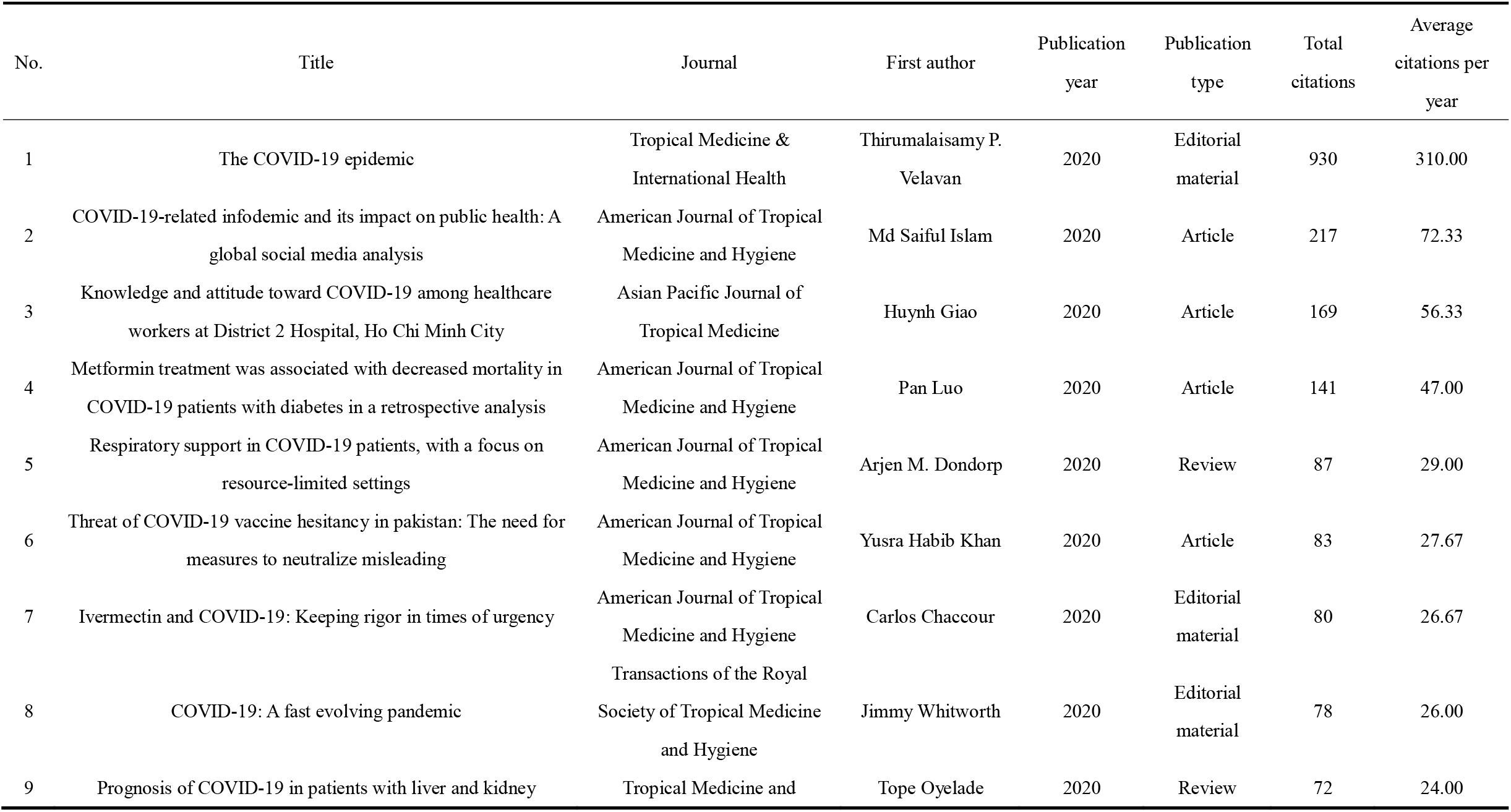
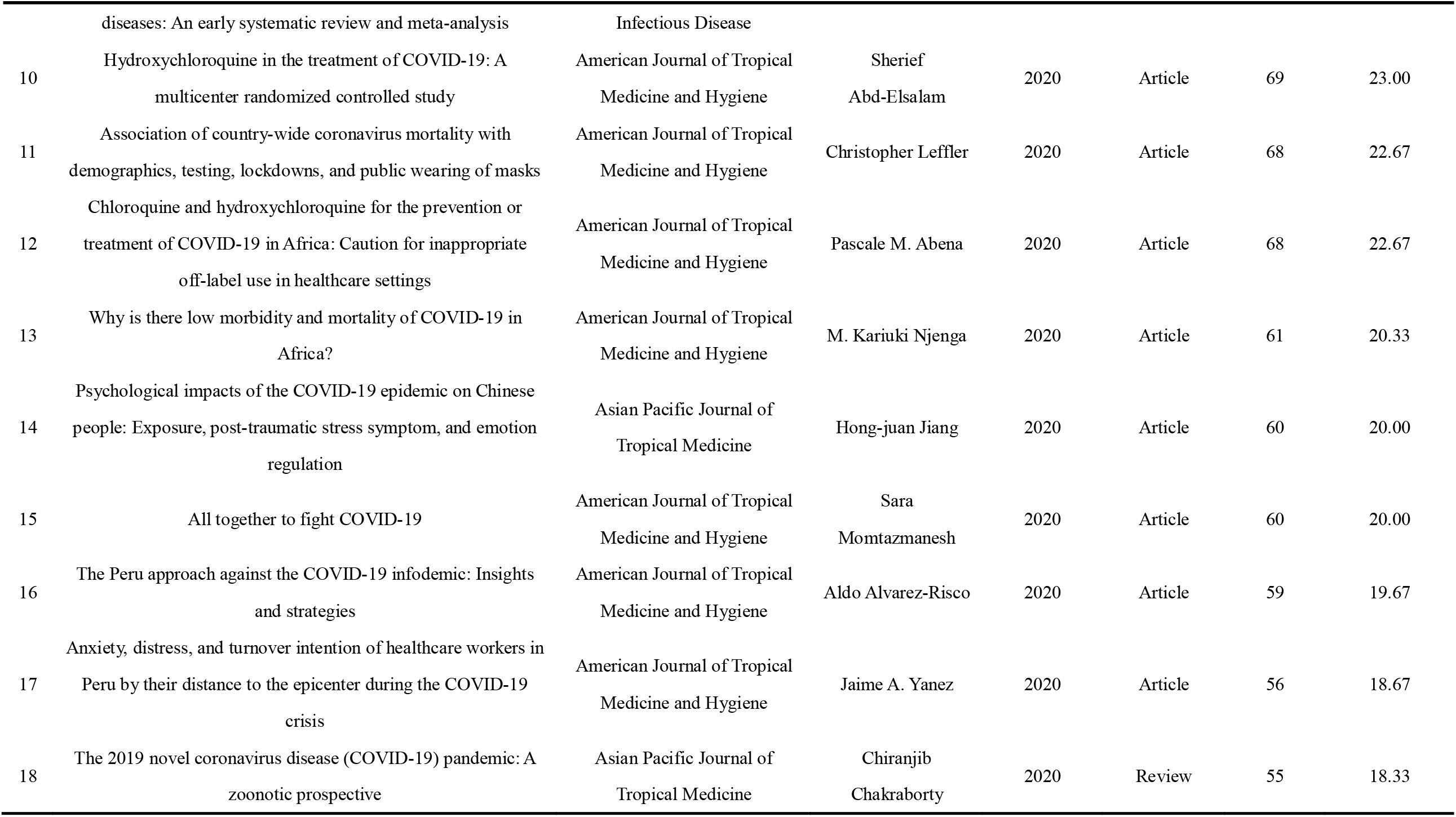
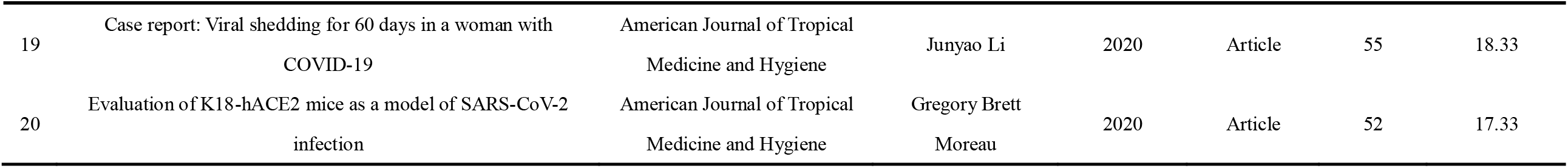
Top-20 most cited papers.

The citation performance analysis elucidated that the documents of interest were frequently cited, while more reuse of them should be expected. To master the topics discussed in this field, the most cited papers shown in **Table 2** were recommended to be referred to.

### 3.4 Co-authorship

The co-authorship of the 783 documents was assessed on the level of regions, organizations, and authors, from ‘macroscopic’ to ‘microscopic’. The VOSviewer software widely used in knowledge domain visualization was employed henceforth^[36-38]^. In the visualized networks, the diameter of an item represented the strength (publication number) and the boldness of a connecting line was proportional to the link strength (frequency of co-authorship of co-occurrence). Item colors indicated different categories and mean publication times for cluster networking and time-overlay networking, respectively^[39]^.

The visualization of co-authorship, regions is provided in **Figure 4**. With the threshold of 15, 29 regions were displayed on the network. The dense network suggested that the cooperation between regions was quite active. USA, England, India, Switzerland and Italy are located at the center of the network, acting as the collaborative pivots, with 28, 27, 25, 18 and 16 links built, respectively. Among them, the USA, England, and India were the top 3 contributing regions in **Figure 2(a)**, possessing a total link strength of 304, 251 and 103, respectively. The most intense co-authorship tiles included USA-England (link strength = 44), USA-Nigeria (link strength = 29), England-Netherlands (link strength = 21) and England-Switzerland (link strength = 20). It was indicated that the collaboration between developed regions was prevailing and that between tropical and non-tropical regions also played an important part.

**Figure 4.**
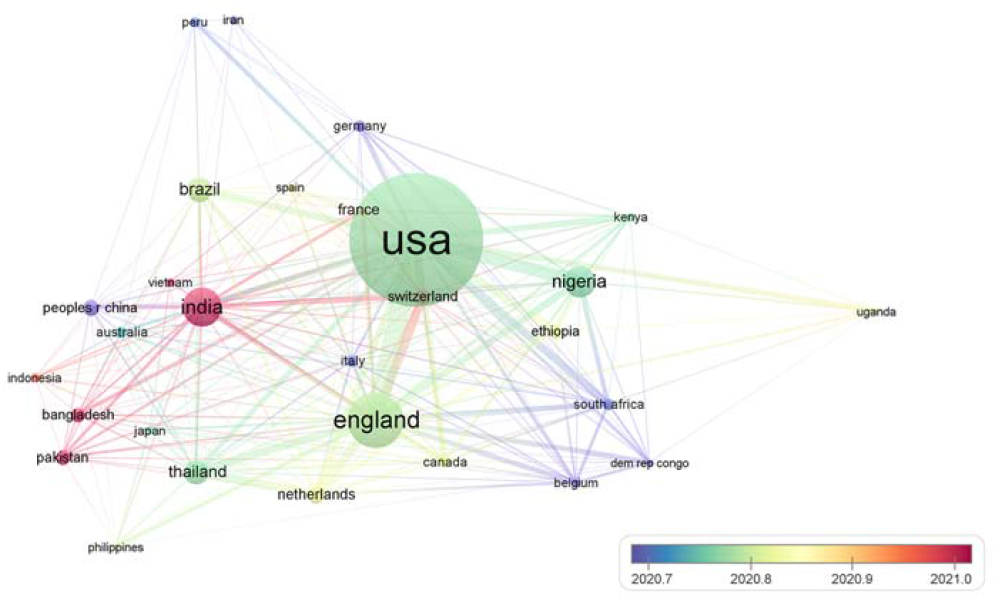
Co-authorship, regions time-overlay visualization.

**Figure 4** also presents the information about mean publication time, as a time-overlay visualization. China, Italy, South Africa, *etc*., marked in purple, launched cooperative research in an early stage (mean publication time ∼2020.7). Marked in green, regions like the USA, England, Nigeria, and Thailand also joined the conversation in an early phase (mean publication time ∼2020.8). The regions like India, Bangladesh, and Pakistan were colored in red (mean publication time ∼2021.0), exhibiting a recent active role in international cooperation.

The visualization of the full data set of co-authorship, organizations are displayed in the upper panel of **Figure 5**. With the threshold of 10, 20 organizations were involved. However, this panel could not introduce much valuable information due to the condensed layout, expect that Hamad Medical Corporation (Qatar) did not cooperate with other organizations. To achieve a better interpretation, the condensed items in the pink box were magnified as the lower panel.

**Figure 5.**
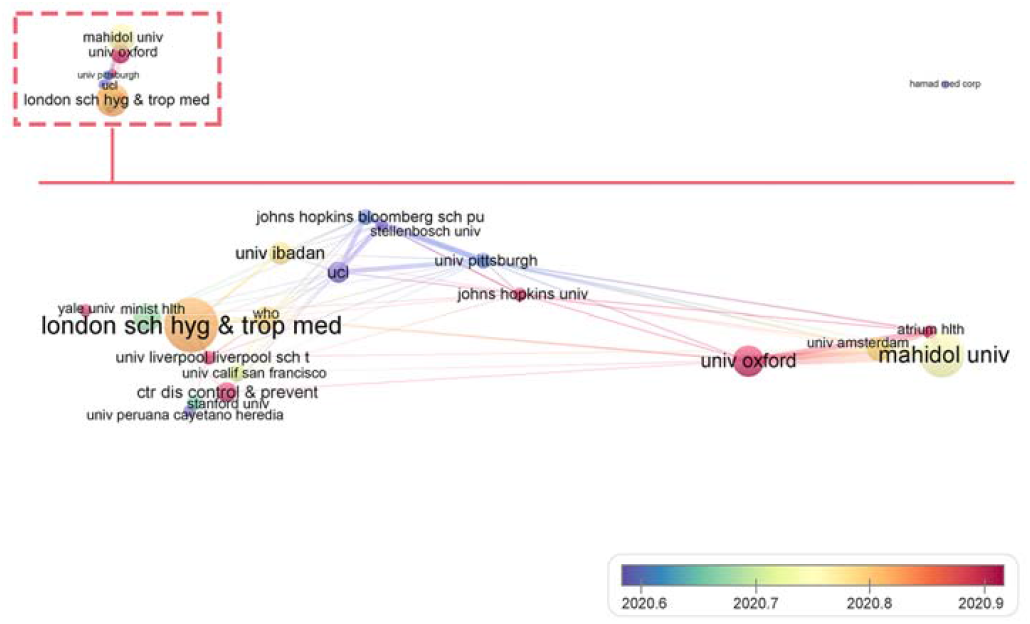
Co-authorship, organizations time-overlay visualization.

The network was of a spindle-like shape, whose density was much lower than in **Figure 4**. University of Pittsburgh (USA), John Hopkins University (USA), and the University of California at San Francisco (USA) were cooperative pivots, establishing 13, 12 and 12 links, respectively. As for the total link strength, Mahidol University (Thailand, 48), University of Oxford (England, 45) and University of Amsterdam (the Netherlands, 41) stood out. University of Oxford-Mahidol University (England-Thailand, 17), Mahidol University-University of Amsterdam (Thailand-Netherlands, 15), and University of Oxford-University of Amsterdam (England-Netherlands, 15) were the most intense cooperative tiles, indicating that collaboration was widely established between non-tropical and topical/non-tropical organizations. However, the collaborative relationship between topical organizations was not evidently exhibited herein.

Johns Hopkins Bloomberg School of Public Health (USA), Stellenbosch University (South Africa), University of Pittsburgh, University College of London (England, UCL), and Universidad Peruana Cayetano Heredia (Peru) participated in the cooperation in a relatively early period (mean publication time ∼2020.6). Organizations like the University of Oxford (England), John Hopkins University (USA), and Atrium Health (USA) were more recent collaborators, with a mean publication time of about 2020.9. Subsequently, the co-authorship profile of authors is illustrated in **Figure 6**. With a threshold of 5, 62 authors were included in the visualization. Overall, the pattern of this profile was very different from **Figure 4** and **Figure 5**. There were a number of individual author groups. To be specific, 16 author groups did not share co-authorship. The seven author groups of John Woodford *et al*. (primary affiliation: National Institute of Allergy and Infectious Diseases, USA), Alimuddin Zumla *et al*. (primary affiliation: University College of London, England), Wafaie W. Fawzi *et al*. (primary affiliation: Harvard University, USA), Anthony D. Harries *et al*. (primary affiliation: International Union Against Tuberculosis and Lung Disease, France), Marcus J. Schultz *et al*. (primary affiliation: Mahidol University, Thailand), Don Eliseo (III) Lucero-prisno *et al*. (primary affiliation: London School of Hygiene & Tropical Medicine, England), and Amber Castillo *et al*. (primary affiliation: The University of New Mexico, USA) were representative large groups.

**Figure 6.**
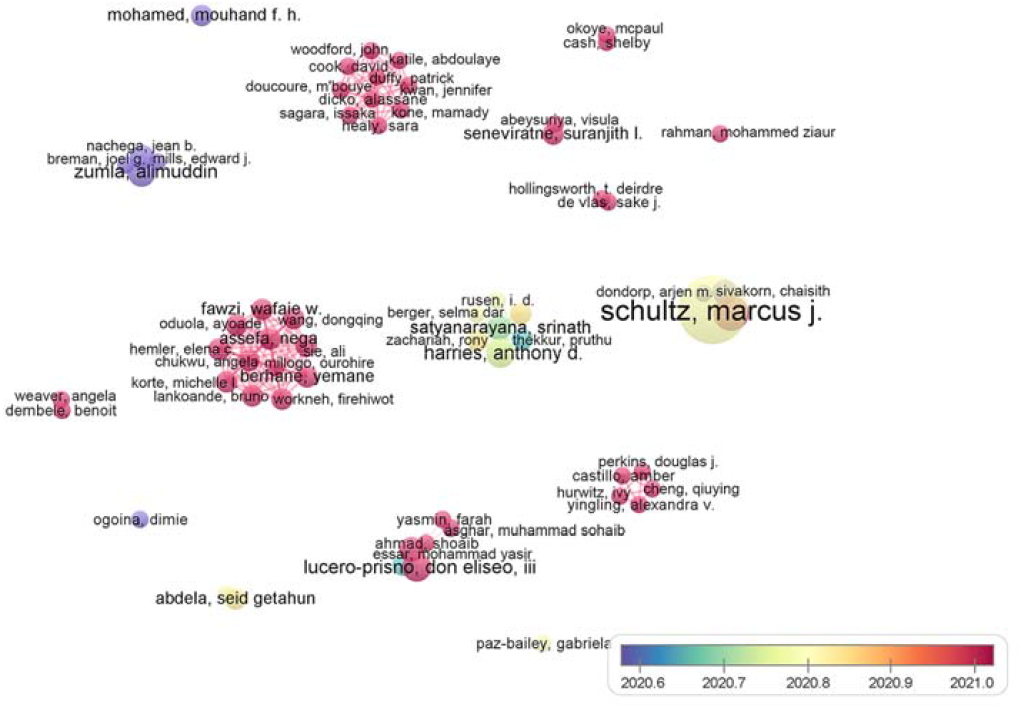
Co-authorship, authors time-overlay visualization.

Seen from the ∼2020.6 mean publication time, the three author groups of Mouhand F.H. Mohamed (primary affiliation: Hamad Medical Corporation, Qatar), Alimuddin Zumla *et al*. (primary affiliation: University College of London, England), and Dimie Ogoina (primary affiliation: Niger Delta University Teaching Hospital, Nigeria) involved in this field in an early stage. More than half of (9/16) author groups in Figure 5 possessed the mean publication time of ∼2021.0, demonstrating that a large proportion of fruitful authors were still active in academic cooperation.

By far, the co-authorship profile of the retrieved documents was analyzed. From regions, organizations to authors, or macroscopic to microscopic, the cooperation network seemed to become looser. It was conjectured that less fruitful organizations or authors that were not involved in **Figure 5** and **Figure 6** had built cooperation relationships, adding to the dense macroscopic (geographical) network. These results also inspired us not to neglect yet to value the contributions from non-top organizations or authors^[40]^.

### 3.5 Co-citation and bibliographic coupling

To describe the state-of-the-art knowledge domain, co-citation and bibliographic coupling analyses were conducted. The definition of co-citation and bibliographic coupling was as follows. Co-citation: Two references not limited to the current set were mutually cited by a publication in the current set; Bibliographic coupling: Two publications in the current sets mutually cited a reference not limited to the current set^[41]^. According to the above definition, the items in co-citation might be older documents published before the COVID-19 epidemic or irrelevant documents published after the epidemic onset, while those in bibliographic coupling belonged to the 783 documents.

**Figure 7(a)** illustrates the co-citation cluster network. The threshold 20 resulted in 11 qualified items. As these papers were frequently referenced by the retrieved documents, it was regarded that they constructed the knowledge base for this field^[42]^. Three clusters colored red, green and blue were shown, representing different elements of the knowledge base.

1. Red cluster, 5 items: Literature (Zhu N, 2020)^[43]^, (Dhama K, 2020)^[44]^, (Wu ZY, 2020)^[45]^, (Zhao JJ, 2020)^[46]^, and (Lee K, 2009). The last item was irrelevant to the COVID-19 pandemic and thus was omitted for analysis. This cluster was mainly about the early knowledge about the characteristics of COVID-19 and the evolving tendency.
2. Green cluster, 4 items: Literature (Zhou F, 2020)^[47]^, (Huang CL, 2020)^[48]^, (Chen NS, 2020)^[49]^, and (Wang DW, 2020)^[50]^. Papers in this cluster mainly dealt with the clinical and epidemiological investigations of COVID-19 in China.
3. Blue cluster, 2 items: Literature (Horby P, 2020)^[51]^, and (Guan Wei-jie, 2020)^[52]^. The former item was a clinical study on the drug dexamethasone administrated to hospitalized COVID-19 patients, while the latter item discussed clinical characteristics of COVID-19 in China, both published in *New England Journal of Medicine*.

**Figure 7.**
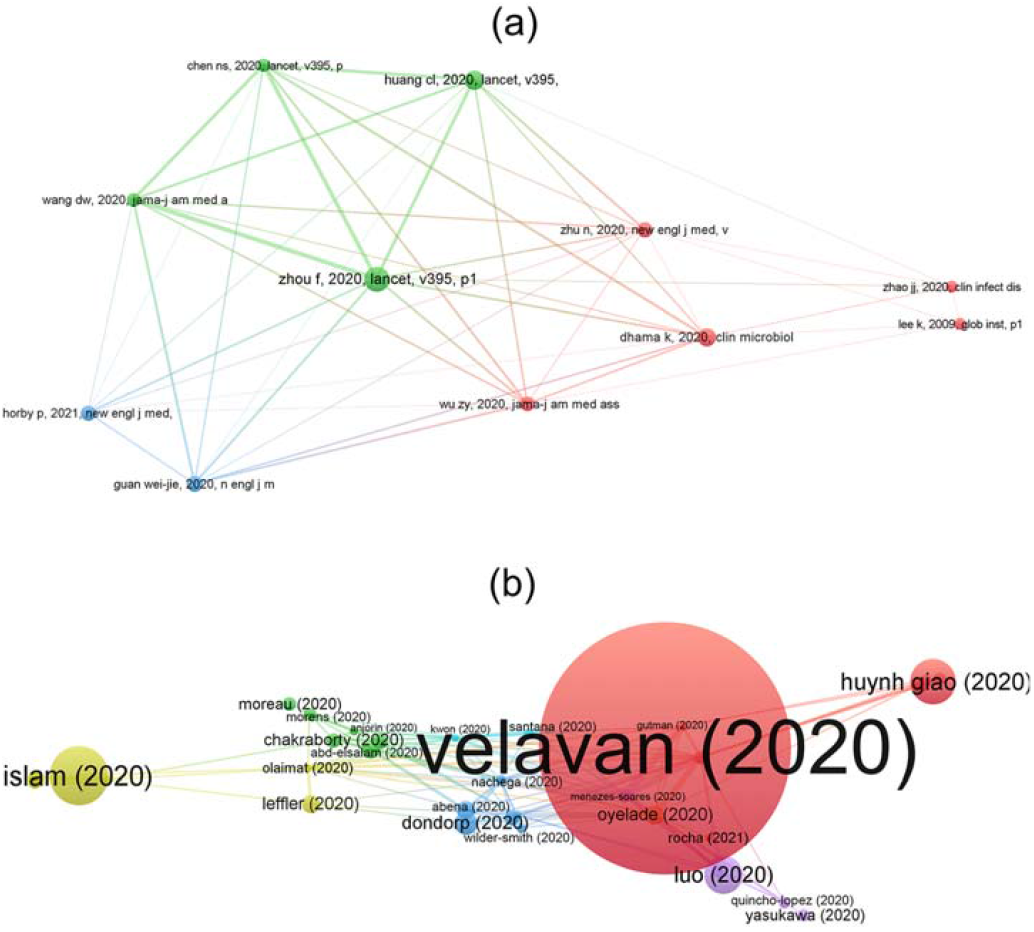
Co-citation cluster network visualization (a) and bibliographic coupling network visualization: partial set of items (b).

The bibliographic coupling network visualization of a full set of items (42) is shown in **Figure S1**. As the seven documents on the right part did not couple with the others, they added no significance to the analysis. Under this circumstance, the condensed cluster on the left part (35 items) was magnified as **Figure 7(b)**, to mine meaningful information. The coupled items, especially in the same cluster, share a high similarity of references, implying that they had similar research intention, protocol or outcome^[53]^. The typical papers in each cluster might be viewed as the classic research paradigm^[54]^. Seven clusters colored in red, green, blue, yellow, purple, cyan and orange were obtained in **Figure 7(b)**.

1. Red cluster, 7 items. Representative literature (Velavan, 2020)^[18]^, and (Huynh Giao, 2020)^[55]^: General knowledge toward COVID-19.
2. Green cluster, 6 items. Representative literature (Chakraborty, 2020)^[56]^, and (Morens, 2020)^[57]^: The origin of COVID-19 from a zoonotic perspective.
3. Blue cluster, 6 items. Representative literature (Nachega, 2020)^[58]^, and (Abena, 2020)^[59]^: Treatment of COVID-19 and its outcome, especially in Africa.
4. Yellow cluster, 6 items. Representative literature (Islam, 2020)^[60]^, and (Alvarez-risco, 2020)^[61]^: The strategies to combat COVID-19 infodemic.
5. Purple cluster, 5 items. Representative literature (Quincho-lopez, 2020)^[62]^, and (Jiang, 2020)^[63]^: The complications of COVID-19.
6. Cyan cluster, 3 items. Representative literature (Faico-Filho, 2020)^[64]^, and (Kwon, 2020)^[65]^: The impact of viral load on the severity of COVID-19.
7. Orange cluster, 2 items. Representative literature (Oyelade, 2020)^[66]^, and (Rocha, 2021)^[67]^: The health threat of COVID-19 combined with other diseases. It was noteworthy that the latter item possessed the newest publication time, representing the burgeoning topics.

Accordingly, the papers reporting the clinical and epidemiological features of COVID-19 (particularly in China) constructed the knowledge base of this aspect. The current research foci included general knowledge, origin, treatment, infodemic, complication and severity determinant of COVID-19, and the consequence of COVID-19 combined with other diseases was also a vital topic. General knowledge of COVID-19 was the overlap of co-citation and bibliographic coupling behavior.

### 3.6 Co-occurrence of keywords and terms

Last but not least, the co-occurrence of keywords and terms was visualized to figure out the hotspots in COVID-19-related studies published in ‘Tropical Medicine’-entitled journals. In this study, all keywords (keywords defined by authors and added by Web of Science) and terms in the title and abstract were analyzed.

The network visualization of 55 keywords’ co-occurrence is shown in **Figure 8. Figure 8(a)** depicts the cluster network, where six clusters were obtained. Except for the search themes (COVID-19, SARS-Cov-2, *etc*.), the typical keywords of each cluster were as follows.

**Figure 8.**
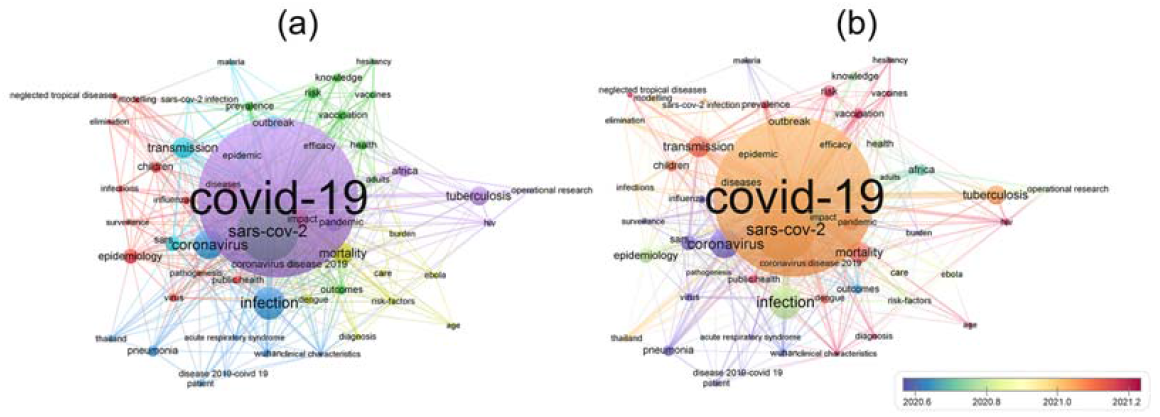
Keywords co-occurrence network visualization: cluster (a) and time-overlay (b).

1. Red cluster, 13 items: About the epidemiology and public health aspect, characterized by ‘epidemiology’, ‘public health’ and ‘surveillance’, *etc*.
2. Green cluster, 13 items: About the aftermath of insufficient vaccination level, characterized by ‘vaccines’, ‘vaccination’ and ‘risk’, *etc*.
3. Blue cluster, 9 items: About the clinical feature, characterized by ‘clinical characteristics’, ‘pneumonia’ and ‘patient’, *etc*.
4. Yellow cluster, 8 items. About the risk in combination with acute infectious diseases, characterized by ‘mortality’, ‘Ebola’ and ‘Dengue’, *etc*.
5. Purple cluster, 6 items. About the risk in combination with chronic infectious diseases, characterized by ‘pandemic’, ‘Human immunodeficiency virus (HIV)’ and ‘tuberculosis’, *etc*.
6. Cyan cluster, 6 items. About the comparison with the SARS epidemic, characterized by ‘SARS’, ‘transmission’ and ‘outbreak’, *etc*.

Taking mean publication time into consideration (∼2020.6), ‘Wuhan’ and ‘pneumonia’ were ‘old’ keywords. This was because in the early stage of the epidemic, the disease had not been officially named, and it was initially considered a type of pneumonia that originated from Wuhan, China (so-called ‘Wuhan Pneumonia’). However, after the knowledge accumulated about the disease, the inappropriate code with suspicion of stigmatization was prohibited^[68]^. In addition, the epidemic rapidly spread outside the Wuhan region. As a result, the heat of ‘Wuhan’ and ‘pneumonia’ dropped in a later period. With the mean publication time ∼2021.2, the keywords like ‘clinical characteristics’, ‘diagnosis’, ‘vaccines’, ‘vaccination’, ‘hesitancy’, ‘HIV’, and ‘neglected tropical diseases’ were the recent hot keywords. The diagnosis and clinical characteristics of COVID-19 were still emphasized, and the vaccination hesitancy and the combination with other diseases were newly burgeoning issues.

Terms co-occurrence are considered non-keyword terms in publications and might be critical addenda to the keywords co-occurrence analysis^[69]^. The results (54 terms) are visualized in **Figure 9**, where predominant terms were identical to or derived from keywords in **Figure 8**. For example, ‘COVID’, ‘SARS CoV’, ‘pandemic’, ‘infection’, ‘patient’, ‘Africa’, ‘mortality’, ‘outcome’, ‘diagnosis’, ‘transmission’, ‘pneumonia’, and so forth. Some protocol-related new terms emerged, like ‘participant’, ‘treatment’, ‘day’ and ‘year’, which probably stemmed from clinical reports. It was worth mentioning that ‘vaccine’ and ‘vaccination’ were also hot terms in **Figure 9 (b)**, mirroring the heated discussion about COVID-19 vaccines.

**Figure 9.**
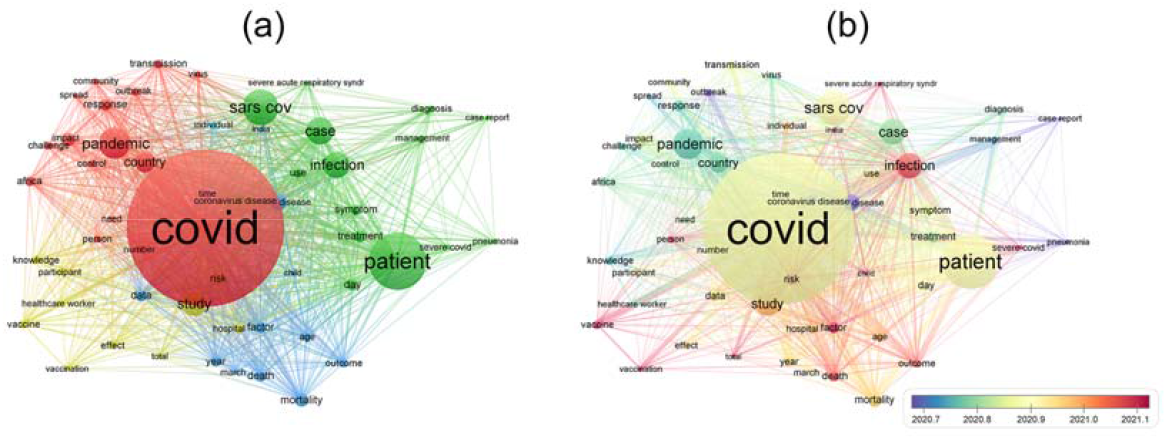
Keywords co-occurrence network visualization: cluster (a) and time-overlay (b).

The co-occurrence analysis basically revealed the current and emerging hotspots in this field. Nevertheless, to ensure the readability and resolution of the figure, a certain threshold was set for visualization. It should be admitted that some extremely new trends introduced by a small number of papers would have been ignored. The authors will attempt to analyze these new tendencies in their future work, with the help of other tools like Python^[70]^.

## 4 Discussion

Having scrutinized the document overview, basic bibliometric characteristics, citation performance, co-authorship, co-citation, bibliographic coupling, and co-occurrence of keywords and terms, the current status and emerging trends of this field were unveiled.

### 4.1 Current status

1. Bibliometric perspective: The topic had turned into a global concern, and investigators outside topical regions made great contributions. Article, meeting abstract and editorial material were the predominant publication type, while review only accounted for a small part. More reviews should be contributed to claim the progress on COVID-19 knowledge. Some ‘Tropical Medicine’-entitled journals publish a small number of relevant papers, and they were suggested to attract more related submissions. Besides, the citation performance could be improved, and attention from other research areas was demanded.
2. Research topics: The clinical features of COVID-19 patients were the mainstream topic. The origin, treatment, infodemic, complication and severity determinants of COVID-19 also provoked intensive discussions.

### 4.2 Emerging trends

1. Bibliometric perspective: Revealed by the co-authorship profile, regions like India, Bangladesh and Pakistan, organizations like the University of Oxford, John Hopkins University and Atrium Health and authors like Wafaie W. Fawzi, John Woodford, and Don Eliseo (III) Lucero-prisno were the recently active contributors. To seek collaboration with them was a beneficial choice for emerging investigators in this field.
2. Research topics: The hesitancy in making vaccines against SARS-CoV-2 and the circumstance where COVID-19 coexisted with other tropical diseases were two recent research hotspots. From the authors’ point of view, more efforts should be made to investigate these two issues, to achieve better epidemic management in tropical regions.

## 5 Conclusion

In this work, the current status and emerging trends of COVID-19-related studies in seven ‘Tropical Medicine’-entitled journals were analyzed from a bibliometric perspective. First, the result of the document overview demonstrates a relatively high level of OA publication. The basic bibliometric characteristics were then evaluated. The results show that to publish COVID-19-related works in ‘Tropical Medicine’-entitled journals was a global trend and studies on this topic have provoked international concern and discussion. The citation performance analysis revealed that more reuse of publications should be expected. The collaboration between developed regions was prevailing, and that between tropical and non-tropical regions also played an important part. The results of co-citation and bibliographic coupling analyses showed that studies on the general knowledge, origin, treatment, infodemic, complication, and severity determinant of COVID-19 served as the current research foci for this field and that the consequence of COVID-19 combined with other diseases was also a vital topic. Hotspot mining was achieved through the analysis of keyword co-occurrence and terms. It was revealed that the keywords like ‘clinical characteristics’, ‘diagnosis’, ‘vaccines’, ‘vaccination’, ‘hesitancy’, ‘ HIV’, and ‘neglected tropical diseases’ were the current hot words, and the vaccination hesitancy and the combination with other diseases were newly burgeoning issues. Overall, this work described a whole bibliometric picture of COVID-19-related studies in seven ‘Tropical Medicine’-entitled journals.

## Data Availability

All data produced in the present work are contained in the manuscript.

## 6 Supplementary materials

Supplementary material: Tables and figures about research areas, top funding agencies and bibliographic coupling visualization.

## 8 Acknowledgment

We thanked the financial support from the National Natural Science Foundation of China, under grant no. 82104070 and 82003665, and from the Guangzhou Science and Technology Plan Project, under grant no. 202201010589.

