## Supplementary material for "Current Status and Emerging Trends of COVID-19-related Studies in Seven ‘Tropical Medicine’-entitled Journals": Table S1,Table S2&Figure S1

**Table S1** Research areas of the retrieved documents.

| No. | Research area | Count | Percentage |
| --- | --- | --- | --- |
| 1 | Tropical Medicine | 783 | 100.00 |
| 2 | Public, Environmental & Occupational Health | 716 | 91.44 |
| 3 | Infectious Diseases | 79 | 10.09 |
| 4 | Parasitology | 67 | 8.56 |

**Table S2** Top-5 funding agencies of the retrieved documents.

| No. | Funding agency | Count | Percentage |
| --- | --- | --- | --- |
| 1 | United States Department of Health Human Services | 34 | 4.34 |
| 2 | National Institutes of Health | 31 | 3.96 |
| 3 | Wellcome Trust | 15 | 1.92 |
| 4 | Bill Melinda Gates Foundation | 14 | 1.79 |
| 5 | CGIAR | 12 | 1.53 |
| 5 | Conselho Nacional de Desenvolvimento Cientifico e Tecnologico | 12 | 1.53 |

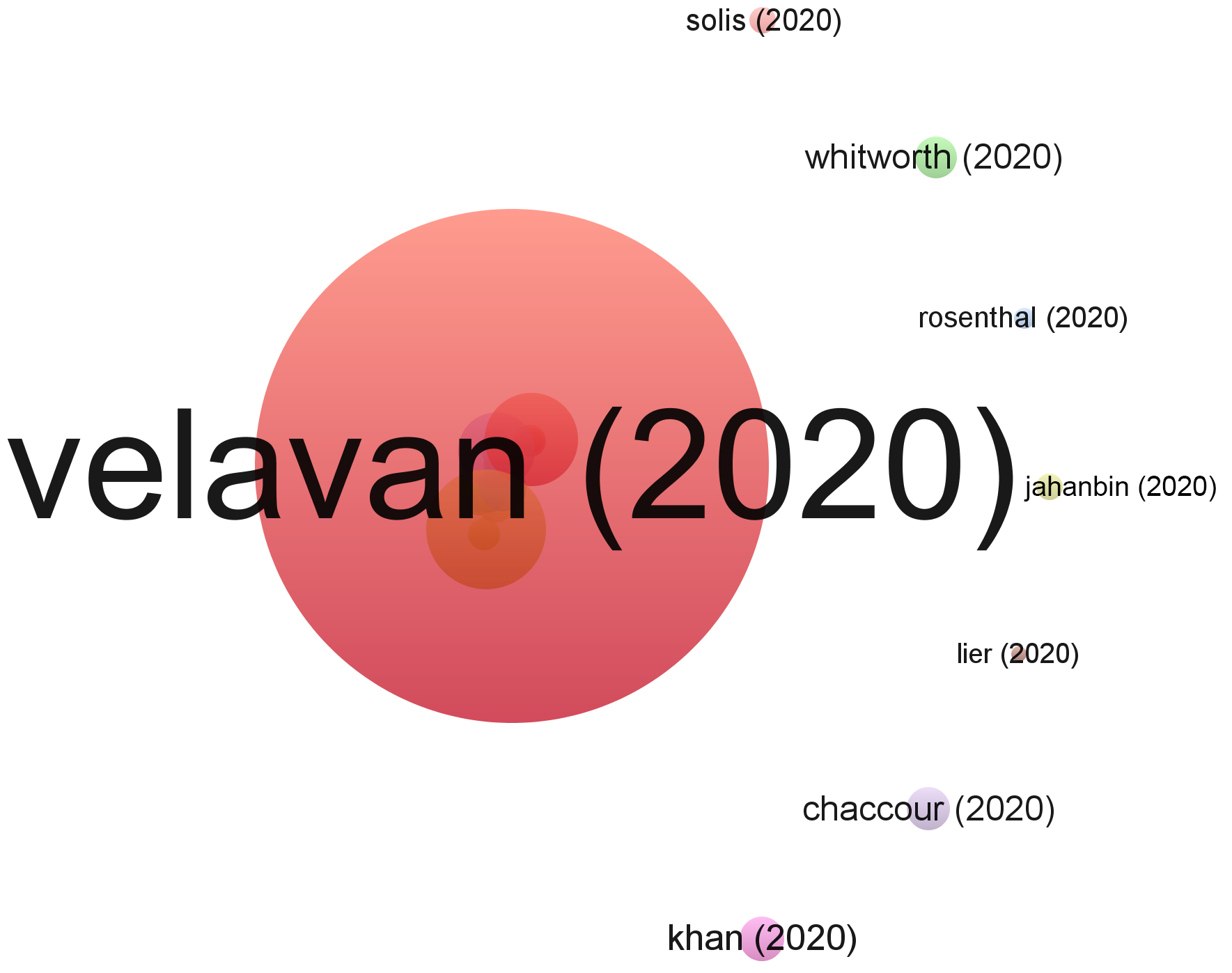

**Figure S1** Bibliographic coupling network visualization: full set of items.
